# Heterogenous treatment effects of blood transfusion in hospitalized patients with congestive heart failure

**DOI:** 10.64898/2026.03.13.26348365

**Authors:** Nicholas A. Bosch, Anica C. Law, Allan J. Walkey

## Abstract

**Background:** Anemia is nearly ubiquitous in hospitalized patients with congestive heart failure (CHF), yet little data informs the decision to transfuse blood in this population.

**Objectives:** To determine average and heterogenous effects of blood transfusion in hospitalized patients with CHF.

**Methods:** We performed a multicenter retrospective cohort study with individual treatment effect analysis using the Premier Healthcare Database (2022-2024). Adult patients with CHF hemoglobin concentrations between 6.5-7.4 g/dL were included. The exposure was blood transfusion based on hemoglobin concentration threshold of <7.0 g/dL. The primary outcome was hospital free days by day 28 (HFDs). We determined the average effect of transfusion using instrumental variable analysis based on a hemoglobin threshold of 7.0 g/dL and estimated the predicted effects of transfusions on HFDs (i.e., conditional average treatment effects) for individual patients using causal forest machine learning models.

**Results:** We included 31,408 patients in a derivation cohort and 30,677 in a validation cohort, of which 13,437 (42.8%) and 13,334 (43.5%) received transfusions, respectively. The average association between transfusion and HFDs suggested harm (derivation mean difference −1.8 [95% CI −2.3, −1.3] days; validation mean difference −1.5 [95% CI −2.0, −1.0] days). However, the effects of transfusion were heterogenous (p=0.001) with the strongest drivers of transfusion benefit being transfusion on hospital day 1 and low serum bicarbonate concentration.

**Conclusions:** The average association between transfusion and HFDs in hospitalized patients with CHF suggested harm; however, there were potential small benefits early in hospitalization and in those with low serum bicarbonate concentrations.

**Key Points:** *Question:* Among hospitalized patients with congestive heart failure and anemia, are the effects of packed red blood cell transfusion heterogeneous based on readily measured clinical characteristics?

*Findings:* In this multicenter retrospective cohort study, associations between packed red blood cell transfusion versus no transfusion on hospital free days by day 28 suggested harm on average but were heterogenous across machine-learning derived deciles of predicted transfusion effect. The strongest drivers of packed red blood cell transfusion benefit were transfusion earlier in hospitalization and low serum bicarbonate.

*Meaning:* Among hospitalized patients with congestive heart failure and anemia, the effects of packed red blood cell transfusion differ between patients based on underlying clinical context.

## Introduction

Nearly 70% of patients with congestive heart failure (CHF) experience anemia and anemia is an independent predictor of poor prognosis among patients with CHF.^1^ In hospitalized settings, clinicians typically administer packed red blood (pRBC) transfusions to treat severe anemia when blood hemoglobin concentrations decrease below 7.0 g/dL with the intent of improving tissue perfusion, including for patients with CHF. However, prior evidence suggests that the average effect of this routine, threshold-based pRBC transfusion practice is either non-beneficial or harmful, and that pRBC transfusion benefits may depend on underlying clinical characteristics.^2,3^ For patients specifically with CHF, risks of pRBC transfusion (e.g., volume overload) and benefits (e.g., improved oxygenation of the myocardium) may be increased compared to patients without CHF. However, neither the average effects of pRBC transfusion nor heterogeneity in the effects of pRBC transfusion on outcomes have been elucidated in patients with CHF but are necessary to inform optimal clinical decision making.

Therefore, in this study we leveraged the quasi-randomization inherent in hemoglobin concentration threshold-based pRBC transfusion and advanced causal inference methods to identify heterogeneous treatment effects of pRBC transfusion among patients hospitalized with heart failure and delineate patient subgroups who may differentially benefit from transfusion.

## Methods

### Cohort

We used the Premier Healthcare Database (PHD) to identify patients for study inclusion. PHD is a multicenter database including hospitalizations from over 1000 United States hospitals.^4^ PHD contains demographics and International Classification of Diseases, Tenth Revision (ICD-10) diagnosis and procedure codes. It also contains hospital-day indexed charge data, which allows sensitive and specific ascertainment of medications and treatment administrations such as pRBC transfusions. A ∼15% subset of hospitals in PHD contribute laboratory data to the database that were used to conduct this study.

We included adult hospitalized patients who were discharged from a PHD hospital between October 1, 2022, and September 30, 2024 who had an ICD-10 diagnosis of congestive heart failure or cardiomyopathy that was coded as being present on admission (eTable 1). Heart failure codes included those that were listed as acute or chronic. We limited to those patients with at least one hemoglobin concentration result between 6.5 and 7.4 g/dL within 28 days of admission because the acceptable variation in laboratory measured hemoglobin concentration is up to ∼0.5 g/dL^5^; thus, over this narrow range and because of the practice of routine threshold-based pRBC transfusion at a hemoglobin concentration of 7.0 g/dL (eFigure 1), patients with hemoglobin concentrations below 7.0 g/dL can be conceptualized as being pseudorandomized to pRBC transfusion due to noise in hemoglobin concentration measurement. For patients with multiple hemoglobin concentrations measured per day, we evaluated the lowest hemoglobin concentration for inclusion in the study. We defined study day 0 as the day that a patient met the hemoglobin concentration criteria. Among patients meeting inclusion criteria on multiple days, one day was randomly selected.

### Exposure and instrumental variable

The exposure of interest was pRBC transfusion on study day 0, which was ascertained using hospital charge codes consistent with our prior work.^2,3^ Because some patients may receive pRBC transfusion for alternative indications (e.g., tissue hypoxia) that may confound the association between pRBC transfusion and outcomes, and that may not be measurable, we opted for an instrumental variable (IV) analysis approach to minimize confounding by indication. Specifically, we used hemoglobin concentration of <7.0 g/dL as an instrument for pRBC transfusion, leveraging its quasi-random assignment effect on transfusion decisions within the defined narrow hemoglobin concentration range.^6^ eFigure 2 shows the proposed relationship between pRBC transfusion, the instrument, confounders, and the outcome. Of importance, IV estimates are only valid among compliers; that is, patients who receive pRBC transfusion for hemoglobin concentrations <7.0 g/dL and do not receive pRBC transfusion for hemoglobin concentrations ≥7.0 g/dL.

### Outcome

The outcome was hospital-free days by day 28 (HFDs), a composite measure of hospital length of stay and all-cause mortality.^7^ HFDs were calculated as 28 minus the number of days from study day 0 to hospital discharge. Patients who died or were discharged to hospice within 28 days or whose days to discharge was longer than 28 days were assigned 0 HFDs. HFDs are patient centered^7^ and encompass the likely downstream benefits (lower mortality and shorter lengths of stay) and risks (higher mortality and longer lengths of stay) of pRBC transfusion. We selected HFDs as the outcome instead of all-cause mortality alone for two main reasons: (1) a signal for differences in all-cause mortality due to a single transfusion event is likely difficult to detect even in large datasets and (2) given a set sample size, continuous measures provide greater statistical power than dichotomous outcomes,^8^ key to generating precise estimates of heterogeneity of treatment effect (HTE) in multi-dimensional data. In this study, our goal was to estimate HTE for pRBC transfusion on HFDs, conditional on patient clinical characteristics. Thus, mean differences in HFDs between pRBC transfusion and no transfusion could potentially range from −28 days (i.e., pRBC transfusion very beneficial) to +28 days (pRBC transfusion very harmful) enabling nuanced quantification of the predicted degree of pRBC transfusion effectiveness.

### Potential contributors to pRBC transfusion heterogeneity of treatment effect

We considered the following clinical characteristics in models as possible contributors to pRBC transfusion HTE: age, sex, days from admission to study day 0, acute organ dysfunctions present on admission,^9^ comorbid diagnoses present on admission,^10,11^ myocardial infarction diagnosis present at admission, acute or acute on chronic CHF present on admission (versus only chronic CHF), complete blood count measurements, metabolic panel measurements, parenteral loop diuretic use, invasive mechanical ventilation use, renal replacement therapy use and care location all on study day 0. Additional definitions are shown in eTable 1.

### Statistical Analysis

The potential outcomes framework^12^ defines individual treatment effects as the difference in outcomes for a single patient between a world where they receive treatment (pRBC transfusion) and a separate parallel world where they do not receive treatment. Individual treatment effects are a theoretical construct that, if known, would allow clinicians to always select the best course of action for every patient. In reality, we only observe a patient receiving or not receiving pRBC transfusion and thus determination of individual treatment effects is not possible. However, methodological advances^13^ have enabled individual treatment effects to be estimated. These estimates are called conditional average treatment effects (CATEs) and are interpreted as the estimated association between pRBC transfusion versus no pRBC transfusion on HFDs in a group of patients with a set of observed characteristics.^14^ Effect estimates from clinical trial subgroup analyses are examples of simple CATEs as they quantify the effect of an intervention on an outcome stratified by a single characteristic (e.g., sex). In this study, we estimated CATEs using an entire set of clinical characteristics to reveal treatment effect heterogeneity based on multi-dimensional clinical data.^14^ CATEs were reported on the mean difference scale. Thus, positive CATEs implied beneficial effects of pRBC transfusion and negative CATEs implied harmful effects of pRBC transfusion.

We split patients into temporally distinct derivation and validation cohorts based on discharge date (derivation: discharge on or before August 31, 2023; validation discharge after August 31, 2023). In both derivation and validation cohorts, we summarized clinical characteristics using counts (percentages) and medians (interquartile ranges [IQRs]). We assessed similarity between clinical characteristics among patients with hemoglobin concentrations 6.5-6.9 g/dL or 7.0-7.4 g/dL using absolute standardized mean differences (SMDs) with SMDs <0.2 defining similarity. We performed standard two-stage least squares (2SLS) IV regression to estimate the population effect of pRBC transfusion on outcomes in each cohort.^15,16^ 2SLS pRBC transfusion effects were reported on the mean difference scale with cluster robust and heteroskedastic consistent standard errors and 95% confidence intervals (CIs) to account for clustering at the hospital-level. We calculated the first-stage F-statistic as a measure of instrument strength and conducted the Durbin-Wu-Hausman test to assess the endogeneity of pRBC transfusion when not considering the instrument.^17,18^

Using the derivation cohort, we trained instrumental variable causal forest models^19^ to estimate CATEs for pRBC transfusion versus no pRBC transfusion on HFDs based on clinical characteristics. The instrumental causal forest algorithm recursively builds an ensemble of decision trees that are split based on clinical characteristics that maximize differences in the estimated CATEs between nodes. Derivation model inputs included the full set of clinical characteristics, the exposure, the instrument, the outcome, and a hospital indicator variable to account for clustering in the model training process.

We applied the fitted instrumental forest model to the validation cohort to generate estimated CATEs for each patient. We tested for heterogeneity in pRBC effectiveness in the validation cohort by including the estimated CATEs as a main effect term and interaction terms with both pRBC transfusion (interaction of interest) and instrument in a standard 2SLS IV model. We then divided the validation cohort into deciles based on estimated CATEs and used 2SLS to generate IV effect estimates within each decile. In exploratory analyses we tested if estimated CATEs identified heterogenous effects of pRBC transfusion for hospital length of stay and all-cause death or discharge to hospice. We used Pearson’s correlation to assess calibration between median estimated CATEs per decile and IV effect estimates from 2SLS. We used Shapley Additive exPlanations (SHAP) analysis^20^ to explore relationships between clinical characteristics and benefits or harm of pRBC transfusion in the validation cohort.

Alpha was set at 0.05. Additional methods are included in the eMethods; analytic code was previously posted online.^21^ This study was deemed not human subjects research (Boston University Institutional Review Board H-44912). Reporting of this study adhered to The Strengthening the Reporting of Observational Studies in Epidemiology Statement (Supplement).^22^

## Results

Among 1,533,586 adult patients admitted to PHD hospitals that contributed laboratory data, a total of 328,006 (21.4%) had CHF present on admission, and 62,085 (4.0%) had at least one hemoglobin concentration between 6.5 and 7.4 g/dL within 28 days of admission and were thus included in analyses (eFigure 2); after splitting there were 31,408 patients in the derivation cohort and 30,677 in the validation cohort. A total of 10,023 (31.9%) and 9,862 (32.1%) had hemoglobin concentrations below 7.0 g/dL and 13,437 (42.8%) and 13,334 (43.5%) received pRBC transfusions in the derivation and validation cohorts, respectively. Clinical characteristics of patients in both cohorts stratified by the hemoglobin level are shown in Table 1; all absolute standardized differences were <0.2, suggesting similarity of cohorts when stratifying by the instrument. A total of 10,180 (32.4%) of patients in the derivation cohort and 9,428 (30.7%) in the validation cohort had a diagnosis of acute CHF (versus only chronic) on admission.

**Table 1:** Clinical characteristics of patients in the derivation and validation cohorts stratified by whether hemoglobin concentration was below 7.0 g/dL.

| Characteristic | Derivation Cohort |  |  |  | Validation Cohort |  |  |  |
| --- | --- | --- | --- | --- | --- | --- | --- | --- |
|  | Overall<br>(n=31,408) | Hemoglobin<br>concentration<br>≥7 g/dL<br>(n=21,385) | Hemoglobin<br>concentration<br><7 g/dL<br>(n=10,023) | SMD | Overall<br>(n=30,677) | Hemoglobin<br>concentration ≥7<br>g/dL (n=20,815) | Hemoglobin<br>concentration<br><7 g/dL<br>(n=9,862) | SMD |
| Age in years, median (IQR) | 73 (63, 81) | 73 (63, 81) | 73 (63, 81) | 0.01 | 73 (64, 81) | 73 (64, 81) | 73 (64, 81) | 0.008 |
| Female sex, No. (%)<sup>c</sup> | 15880 (50.6) | 10785 (50.4) | 5095 (50.8) | 0.008 | 15634 (51.0) | 10573 (50.8) | 5061 (51.3) | 0.01 |
| Days from admission to study<br>day 0, median (IQR) | 3 (1, 7) | 3 (1, 7) | 2 (1, 5) | 0.182 | 3 (1, 7) | 3 (1, 7) | 2 (1, 5) | 0.183 |
| Variables present on admission |  |  |  |  |  |  |  |  |
| NSTEMI or STEMI, No. (%) | 4288 (13.7) | 2921 (13.7) | 1367 (13.6) | 0.001 | 3689 (12.0) | 2477 (11.9) | 1212 (12.3) | 0.012 |
| Acute CHF, No. (%) | 10180 (32.4) | 7130 (33.3) | 3050 (30.4) | 0.062 | 9428 (30.7) | 6530 (31.4) | 2898 (29.4) | 0.043 |
| Acute organ dysfunctions, No. (%) |  |  |  |  |  |  |  |  |
| Cardiac | 7179 (22.9) | 4804 (22.5) | 2375 (23.7) | 0.029 | 7279 (23.7) | 4963 (23.8) | 2316 (23.5) | 0.008 |
| Neurologic | 4681 (14.9) | 3191 (14.9) | 1490 (14.9) | 0.002 | 4522 (14.7) | 3103 (14.9) | 1419 (14.4) | 0.015 |
| Hematologic | 3784 (12.0) | 2493 (11.7) | 1291 (12.9) | 0.037 | 3867 (12.6) | 2618 (12.6) | 1249 (12.7) | 0.003 |
| Hepatic | 655 (2.1) | 411 (1.9) | 244 (2.4) | 0.035 | 618 (2.0) | 392 (1.9) | 226 (2.3) | 0.029 |
| Renal | 13225 (42.1) | 9042 (42.3) | 4183 (41.7) | 0.011 | 12188 (39.7) | 8280 (39.8) | 3908 (39.6) | 0.003 |
| Respiratory | 6589 (21.0) | 4530 (21.2) | 2059 (20.5) | 0.016 | 6188 (20.2) | 4198 (20.2) | 1990 (20.2) | <0.001 |
| Sepsis, No. (%) | 11846 (37.7) | 8195 (38.3) | 3651 (36.4) | 0.039 | 11438 (37.3) | 7923 (38.1) | 3515 (35.6) | 0.05 |
| Comorbidities, No. (%) |  |  |  |  |  |  |  |  |
| Alcohol abuse | 1939 (6.2) | 1281 (6.0) | 658 (6.6) | 0.024 | 1867 (6.1) | 1280 (6.1) | 587 (6.0) | 0.008 |
| Non-metastatic cancer | 4156 (13.2) | 2737 (12.8) | 1419 (14.2) | 0.04 | 4386 (14.3) | 2972 (14.3) | 1414 (14.3) | 0.002 |
| Cardiac arrhythmias | 16826 (53.6) | 11577 (54.1) | 5249 (52.4) | 0.035 | 16743 (54.6) | 11429 (54.9) | 5314 (53.9) | 0.021 |
| Chronic pulmonary disease | 14893 (47.4) | 10191 (47.7) | 4702 (46.9) | 0.015 | 14803 (48.3) | 10172 (48.9) | 4631 (47.0) | 0.038 |
| Coagulopathy | 5669 (18.0) | 3753 (17.5) | 1916 (19.1) | 0.04 | 5752 (18.8) | 3827 (18.4) | 1925 (19.5) | 0.029 |
| Diabetes with complications | 14735 (46.9) | 10182 (47.6) | 4553 (45.4) | 0.044 | 14234 (46.4) | 9747 (46.8) | 4487 (45.5) | 0.027 |
| Nutrition deficiency anemias | 9898 (31.5) | 6904 (32.3) | 2994 (29.9) | 0.052 | 10065 (32.8) | 6999 (33.6) | 3066 (31.1) | 0.054 |
| Dementia | 3083 (9.8) | 2103 (9.8) | 980 (9.8) | 0.002 | 3108 (10.1) | 2088 (10.0) | 1020 (10.3) | 0.01 |
| Fluid and electrolyte disorders | 16842 (53.6) | 11455 (53.6) | 5387 (53.7) | 0.004 | 16752 (54.6) | 11402 (54.8) | 5350 (54.2) | 0.011 |
| Hemiplegia | 606 (1.9) | 431 (2.0) | 175 (1.7) | 0.02 | 622 (2.0) | 429 (2.1) | 193 (2.0) | 0.007 |
| HIV or AIDS | 153 (0.5) | 102 (0.5) | 51 (0.5) | 0.005 | 186 (0.6) | 132 (0.6) | 54 (0.5) | 0.011 |
| Hypertension | 28779 (91.6) | 19663 (91.9) | 9116 (91.0) | 0.036 | 28246 (92.1) | 19180 (92.1) | 9066 (91.9) | 0.008 |
| Liver disease (moderate to severe) | 3692 (11.8) | 2478 (11.6) | 1214 (12.1) | 0.016 | 3694 (12.0) | 2525 (12.1) | 1169 (11.9) | 0.009 |
| Metastatic cancer | 1422 (4.5) | 963 (4.5) | 459 (4.6) | 0.004 | 1501 (4.9) | 1050 (5.0) | 451 (4.6) | 0.022 |
| Peripheral vascular disease | 6909 (22.0) | 4768 (22.3) | 2141 (21.4) | 0.023 | 6689 (21.8) | 4512 (21.7) | 2177 (22.1) | 0.01 |
| Psychiatric disease | 6405 (20.4) | 4391 (20.5) | 2014 (20.1) | 0.011 | 6391 (20.8) | 4474 (21.5) | 1917 (19.4) | 0.051 |
| Pulmonary circulation disorders | 6176 (19.7) | 4325 (20.2) | 1851 (18.5) | 0.044 | 6617 (21.6) | 4534 (21.8) | 2083 (21.1) | 0.016 |
| Renal failure | 19489 (62.1) | 13384 (62.6) | 6105 (60.9) | 0.034 | 19038 (62.1) | 12955 (62.2) | 6083 (61.7) | 0.011 |
| Weight loss | 4556 (14.5) | 3002 (14.0) | 1554 (15.5) | 0.041 | 4391 (14.3) | 2953 (14.2) | 1438 (14.6) | 0.011 |
| Variables on study day 0<sup>a</sup> |  |  |  |  |  |  |  |  |
| White blood count in K/μL, median (IQR)<sup>b</sup> | 8.50 (6.04, 12.03) | 8.40 (6.01, 11.90) | 8.6 (6.1, 12.4) | 0.052 | 8.5 (6, 12.1) | 8.4 (6, 11.9) | 8.6 (6, 12.4) | 0.056 |
| Platelet count in $\times 10^9$/L, median (IQR) | 193 (132, 271) | 195 (134, 272) | 188 (126, 269) | 0.041 | 193 (131, 272) | 195 (132, 274) | 190 (126, 268) | 0.051 |
| Serum sodium concentration (mEq/L), median (IQR) | 138 (135, 141) | 138 (135, 141) | 138 (135, 141) | 0.02 | 138 (135, 141) | 138 (135, 141) | 138 (135, 141) | 0.007 |
| Serum potassium concentration (mEq/L), median (IQR) | 4.1 (3.7, 4.5) | 4.1 (3.7, 4.5) | 4.1 (3.7, 4.6) | 0.056 | 4.1 (3.7, 4.5) | 4.1 (3.7, 4.5) | 4.1 (3.7, 4.6) | 0.061 |
| Serum chloride concentration (mEq/L), median (IQR) | 104 (99, 108) | 104 (99, 108) | 104 (100, 108) | 0.049 | 104 (100, 108) | 104 (100, 108) | 105 (100, 109) | 0.041 |
| Serum bicarbonate concentration (mEq/L), median (IQR) | 24.80 (22, 28) | 25 (22, 28) | 24 (21, 27) | 0.121 | 24 (21, 27) | 24 (21.40, 27.10) | 24 (21, 27) | 0.126 |
| Serum blood urea nitrogen concentration (mg/dL), median (IQR) | 34 (21, 54) | 33 (20, 53) | 35 (21, 56) | 0.056 | 33 (20, 53) | 32 (20, 52) | 33 (21, 54) | 0.071 |
| Glomerular Filtration Rate (mL/min/1.73 m<sup>2</sup>), median (IQR) | 38 (18, 60) | 38 (18, 60) | 38 (18, 60) | 0.008 | 39 (18, 64) | 39.35 (18, 65) | 39 (18, 63) | 0.024 |
| Serum glucose concentration (mg/dL), median (IQR) | 119 (97, 155) | 119 (97, 155) | 119 (98, 156) | 0.018 | 118 (97, 153) | 117 (97, 152) | 119 (98, 156) | 0.042 |
| Parenteral loop diuretic use, | 8100 (25.8) | 5503 (25.7) | 2597 (25.9) | 0.004 | 7422 (24.2) | 5107 (24.5) | 2315 (23.5) | 0.025 |

| No. (%) |  |  |  |  |  |  |  |  |
| --- | --- | --- | --- | --- | --- | --- | --- | --- |
| Antibiotic use, No. (%) | 16999 (54.1) | 11490 (53.7) | 5509 (55.0) | 0.025 | 16548 (53.9) | 11187 (53.7) | 5361 (54.4) | 0.012 |
| Vasopressor use, No. (%) | 3897 (12.4) | 2375 (11.1) | 1522 (15.2) | 0.121 | 3445 (11.2) | 2071 (9.9) | 1374 (13.9) | 0.123 |
| Dobutamine use, No. (%) | 421 (1.3) | 261 (1.2) | 160 (1.6) | 0.032 | 377 (1.2) | 225 (1.1) | 152 (1.5) | 0.04 |
| Milrinone use, No. (%) | 464 (1.5) | 292 (1.4) | 172 (1.7) | 0.028 | 444 (1.4) | 284 (1.4) | 160 (1.6) | 0.021 |
| Invasive mechanical ventilation use, No. (%) | 4373 (13.9) | 2756 (12.9) | 1617 (16.1) | 0.092 | 4061 (13.2) | 2571 (12.4) | 1490 (15.1) | 0.08 |
| Renal replacement therapy use, No. (%) | 3079 (9.8) | 2095 (9.8) | 984 (9.8) | 0.001 | 2950 (9.6) | 1977 (9.5) | 973 (9.9) | 0.012 |
| Intensive care unit location, No. (%) | 7661 (24.4) | 4876 (22.8) | 2785 (27.8) | 0.115 | 6791 (22.1) | 4324 (20.8) | 2467 (25.0) | 0.101 |
| Intermediate care unit location, No. (%) | 5735 (18.3) | 3886 (18.2) | 1849 (18.4) | 0.007 | 5594 (18.2) | 3745 (18.0) | 1849 (18.7) | 0.02 |

<sup>a</sup>Study day 0 defined as the day of hemoglobin concentration measurement.
<sup>b</sup>Variables with missing data (% of patients with missing data): Derivation cohort: white blood cell count (3.7%), platelet concentration (3.9%), sodium concentration (7.7%), potassium concentration (4.5%), chloride concentration (6.5%), bicarbonate concentration (5.4%), blood urea nitrogen concentration (5.0%), creatinine-based glomerular filtration rate (9.3%), glucose (3.7%). Validation cohort: white blood cell count (3.7%), platelet concentration (3.8%), sodium concentration (8.5%), potassium concentration (4.1%), chloride concentration (6.6%), bicarbonate concentration (4.6%), blood urea nitrogen concentration (4.6%), creatinine-based glomerular filtration rate (5.8%), glucose (2.8%).
<sup>c</sup>To protect the identity of the small number of patients in this deidentified study who have sex recorded as “other sex” in the database, sex was dichotomized as female sex or not with patients not specifically identified in the dataset as female sex being classified as not female sex.
SMD: absolute standardized mean difference. HIV: human immunodeficiency virus; AIDS: acquired immunodeficiency syndrome; NSTEMI: Non-ST-Elevation Myocardial Infarction; STEMI: ST-Elevation Myocardial Infarction

### Overall effect of pRBC transfusion

Mean HFDs in the derivation and validation cohorts were 17.6 (standard deviation [SD] 10.1) days and 17.9 (SD 10.0) days, respectively (eTable 2). In both the derivation and validation cohorts, hemoglobin concentration <7.0 g/dL was a strong instrument for pRBC transfusion (F-statistics of 8868 and 9,067, respectively) and the association between pRBC transfusion and HFDs without IV analysis suggested endogeneity (Durbin Wu-Hausman statistics of 99 and 77, p<0.001 and <0.001, respectively). In the derivation cohort, the average association between pRBC transfusion versus no pRBC transfusion and HFDs was −1.8 (95% CI −2.3, −1.3; p<0.001) days, consistent with increased risk from pRBC transfusion. In the validation cohort, the average association between pRBC transfusion versus no pRBC transfusion and HFDs was −1.5 (95% CI −2.0, −1.0; p<0.001) days (eTable 2).

### HTE Derivation cohort

The clinical characteristics used most frequently in tree splits were potassium concentration, blood urea nitrogen, white blood cell count, days from admission to study day 0, and platelet count (Figure 1, eTable 3). In the derivation cohort, the mean estimated CATE (i.e., mean difference in HFDs between pRBC transfusion and no transfusion) was −1.0 (standard deviation 0.9) days, with a minimum of −5.9 and a maximum of +2.6 days across patients (Figure 2).

**Figure 1:**
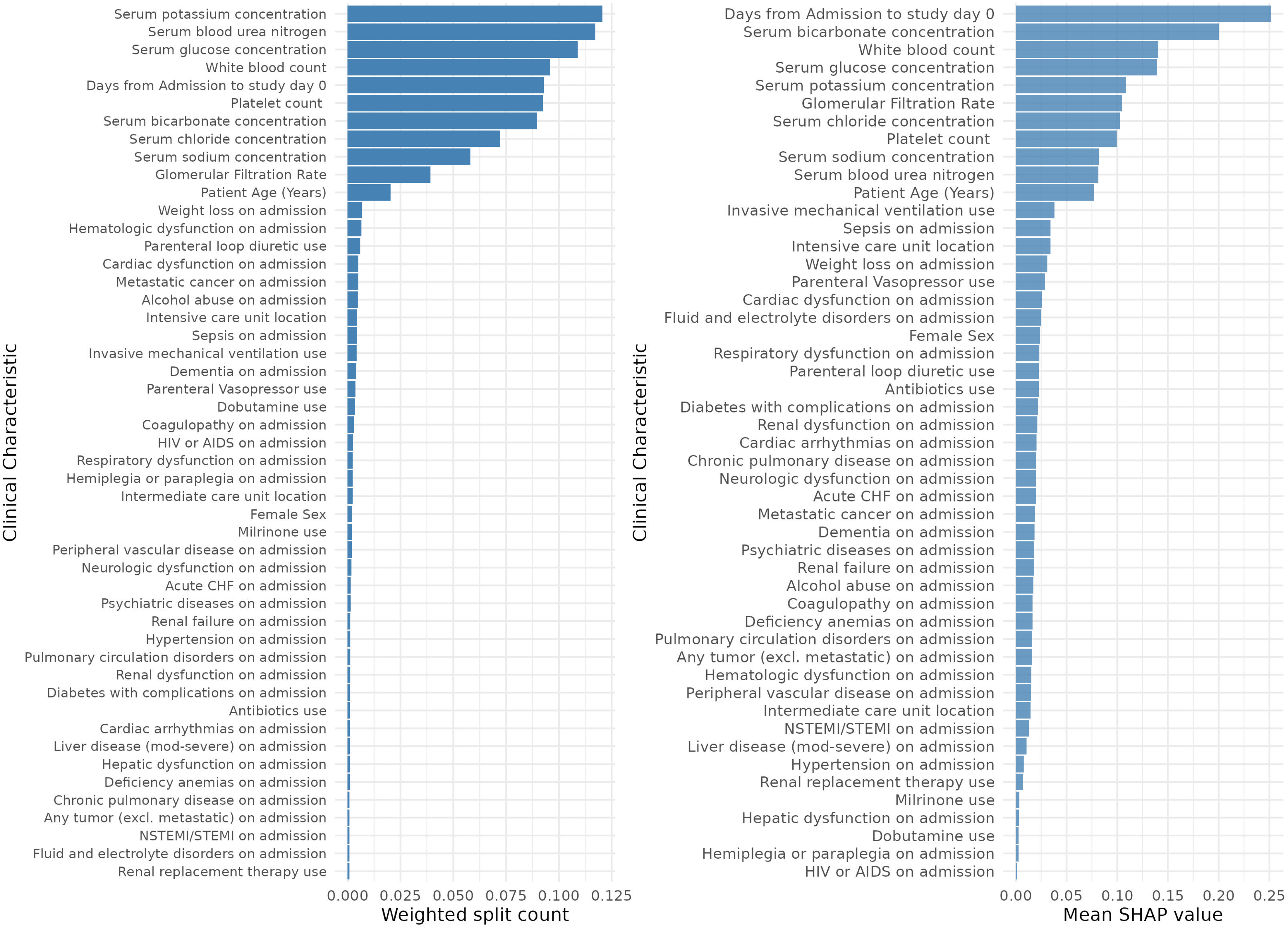
Variable importance Plot in the Derivation and Validation Cohorts. The panel on the left shows the relative importance of each clinical characteristic in the final fitted model in the derivation cohort. Higher weighted splits on the x-axis correspond to that variable being used more frequently in tree splits in the forest model. The panel on the right shows the relative importance of each clinical characteristic based on absolute mean SHAP value in the validation cohort in contributing to heterogenous effects of pRBC transfusion. Variables not reported as “on admission” were measured on study day 0 (i.e., the day of hemoglobin concentration measurement). pRBC: packed red blood cells. SHAP: Shapley Additive exPlanations (SHAP) analysis. BUN: blood urea nitrogen; IMV: invasive mechanical ventilation; ICU: intensive care medicine; HIV: human immunodeficiency virus; NSTEMI/STEMI: non-ST-segment elevation myocardial infraction/ST-segment elevation myocardial infarction; AIDS: acquired immunodeficiency syndrome. WBC: white blood cell count. pRBC: packed red blood cell.

**Figure 2:**
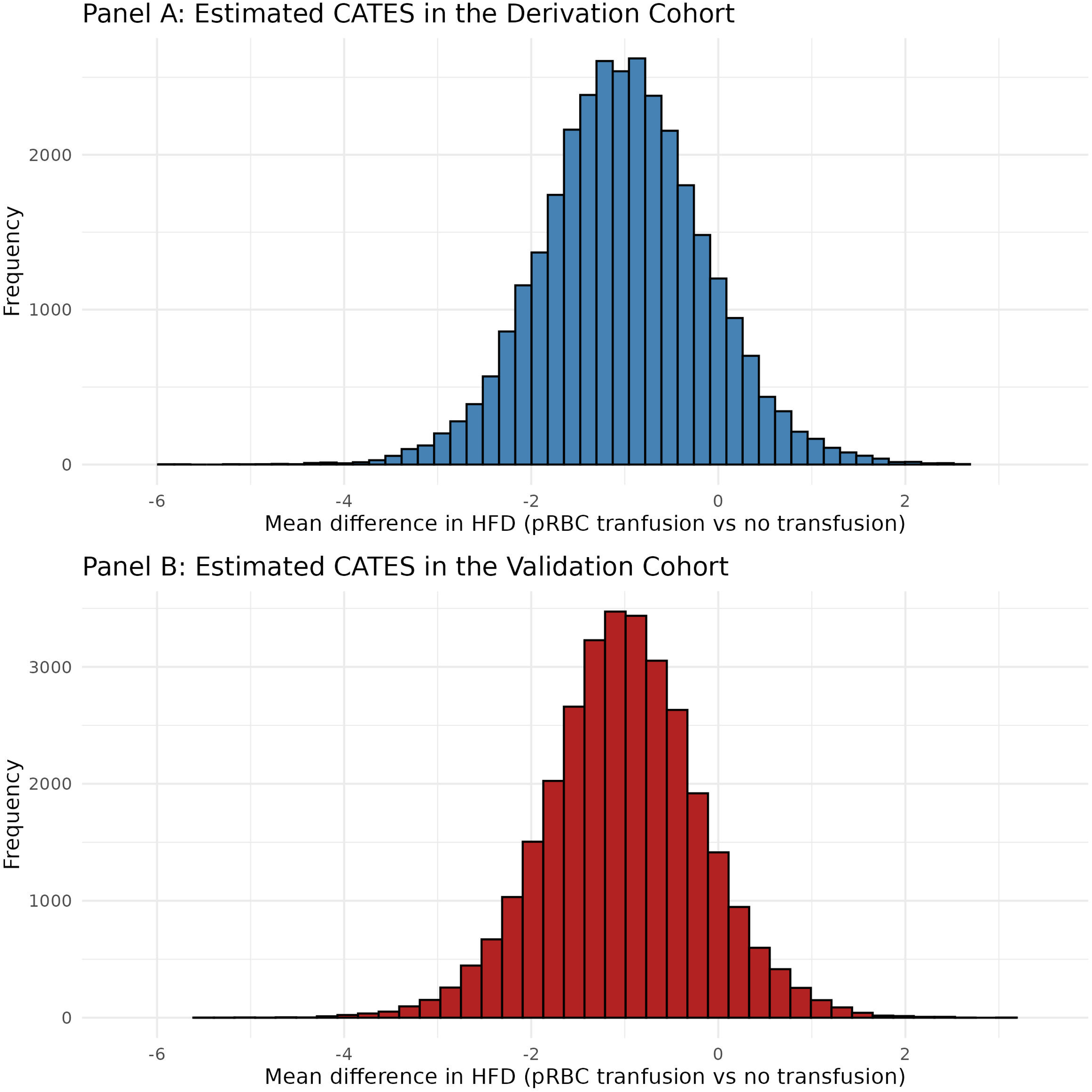
Distribution of estimated CATEs in the derivation and validation cohorts. Shown are the distribution of estimated CATEs across patients in the derivation cohort (from out of bag predictions; top panel) and the validation cohort (bottom panel). CATE: conditional average treatment effects.

### HTE Validation cohort

In the validation cohort, the mean estimated CATE was −1.0 (standard deviation 0.8; minimum −5.6, maximum +3.0) days; only 3,198 (10.4%) patients in the validation cohort had estimated CATEs above 0 suggesting predicted benefit from pRBC transfusion (Figure 2). The 2SLS test for interaction between estimated CATE and pRBC transfusion was significant (p=0.01) implying that the model identified heterogeneous effects of pRBC transfusion on HFDs. However, there were no individual CATE-based deciles in which 2SLS estimates suggested statistically significant benefit of pRBC transfusion (Figure 3). Median estimated CATEs from each decile were well calibrated to 2SLS IV estimates (Pearson Correlation of 0.83 [95% CI 0.43, 0.96], p=0.003; eFigure 3). Clinical characteristics by estimated decile are shown in eTable 4. In exploratory analyses, estimated CATEs were also potentially related to HTE in hospital length of stay (interaction p=0.07); relationships between estimated CATEs and HTE in death or discharge to hospice were less clear interaction p-value 0.17; eFigure 4).

**Figure 3:**
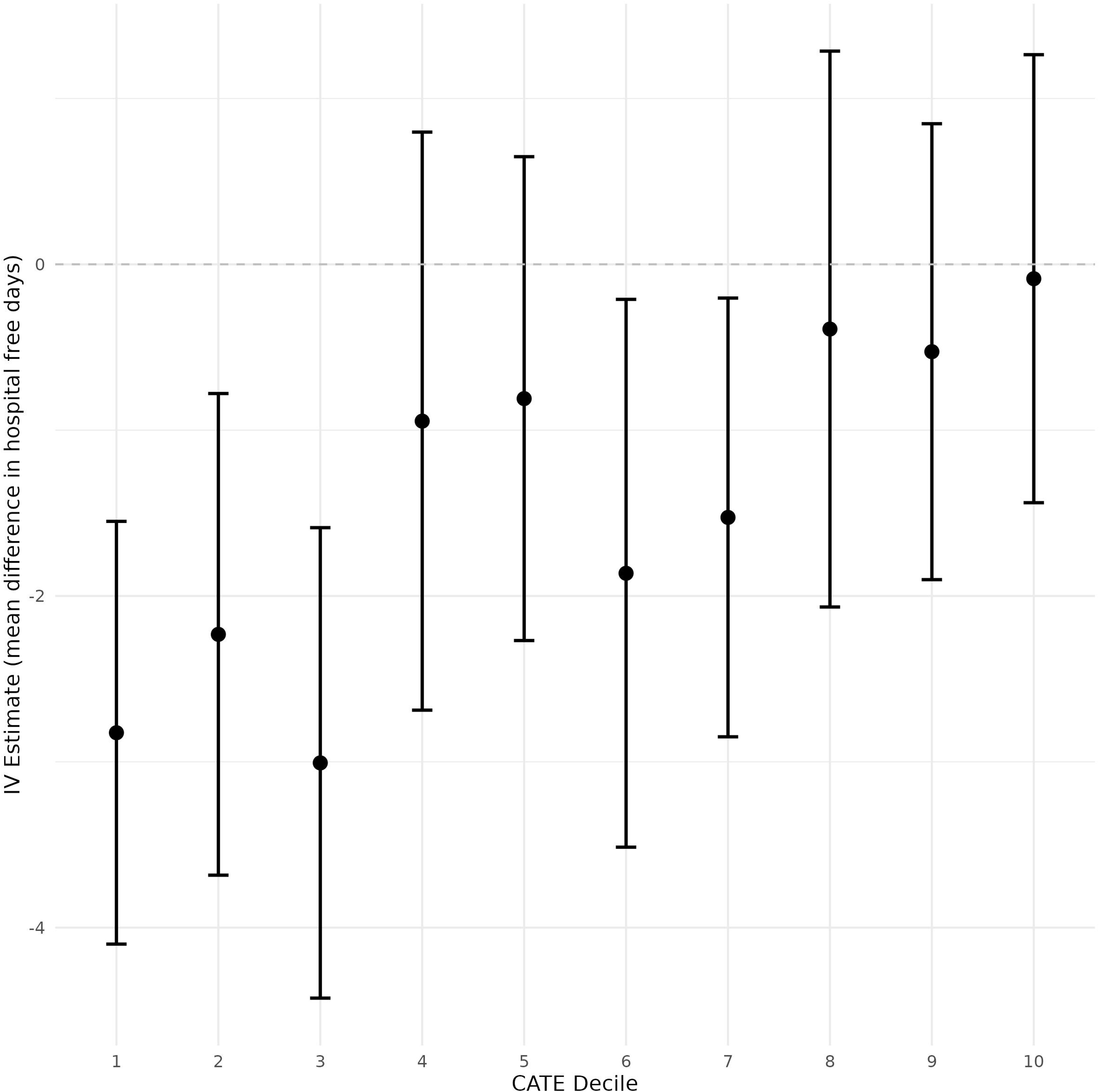
Mean difference in HFD by CATE decile. Shown are two-stage least squares regression estimates of the mean difference in hospital free days by day 28 and robust 95% confidence between pRBC transfusion and no pRBC transfusion for each CATE decile. Negative mean differences in hospital free days correspond to harms from pRBC transfusion and positive mean differences in hospital free days correspond to benefits from pRBC transfusion. CATE: conditional average treatment effect; HFD. Hospital free days by day 2. pRBC: packed red blood cell.

Clinical characteristics that were the strongest drivers of pRBC transfusion treatment effect heterogeneity in the validation cohort were similar to those identified in the derivation cohort, although the specific ordering of relative importance differed (Figure 1). Relationships between values of clinical characteristics and predicted harm versus benefit from pRBC transfusion were complex and non-linear (Figure 3). pRBC transfusion that occurred close to admission, low bicarbonate counts, and low white blood cell counts were associated with positive mean differences in HFDs (i.e., beneficial effects of pRBC transfusion). pRBC transfusion that occured later in the hospitalization, low and high potassium concentrations, and low glomerular filtration rates were associated with the most negative mean differences in HFD (i.e., potential harmful effects of pRBC transfusion) (Figure 4).

**Figure 4:**
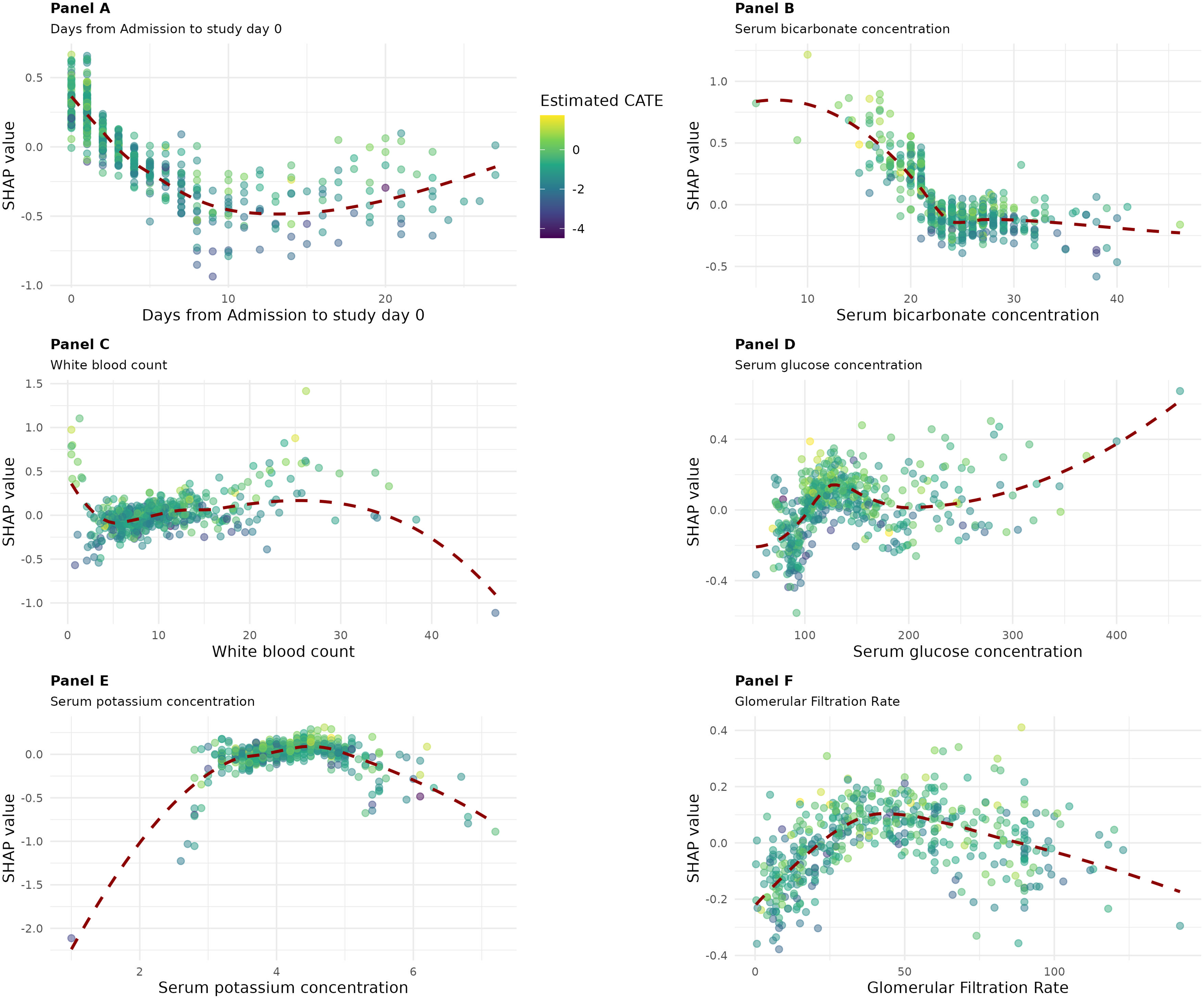
SHAP dependence plots for the six most important clinical characteristics in determining heterogeneous effects of pRBC transfusion. Shown are the relationship between the value of the six most important clinical characteristic (x-axis) for predicting heterogeneous effects of pRBC transfusion and the corresponding SHAP value from 100 randomly selected patients in the validation cohort. SHAP values above 0 correspond to positive HFDs (benefits from pRBC transfusion); SHAP values below 0 correspond to negative HFDs (harms from pRBC transfusion). Each point represents a single patient and is shaded based on estimated CATE for that patient. The dotted red line shows a locally estimated scatterplot smoothing (LOESS) line to visualize the relationship between clinical characteristic and SHAP value. The units for the x-axes are as follows: Panel A: days; Panel B: count x10⁹/L; Panel C: count K/μL; Panel D: mEq/L; Panel E: mL/min/1.73 m²; Panel F: mg/dL. HFDs: hospital free days by day 28. pRBC: packed red blood cells. SHAP: Shapley Additive exPlanations.

## Discussion

In this multicenter retrospective cohort study of adult patients admitted with CHF, we identified heterogeneous effects of pRBC transfusion at a hemoglobin concentration threshold of 7.0 g/dL. Although the average effect of pRBC transfusion suggested harm, instrumental causal forest models predicted that 10% of patients may benefit from pRBC transfusion. The highest benefits of pRBC transfusion were observed when pRBC transfusion was administered early in the hospitalization and in patients with low bicarbonate concentrations, and very low white blood cell counts. However, expected benefits were small with no subgroup of patients showing significant benefit. These results suggest that although most patients with CHF experience harm from pRBC transfusion, effects are heterogenous and can be predicted using routinely collected clinical characteristics that may have value in personalizing pRBC transfusion decisions.

Clinical support tools^23^ and transfusion guidelines^24^ extrapolate data from other contexts to recommend the use of restrictive hemoglobin concentration thresholds (e.g., 7.0 or 8.0 g/dL) over higher thresholds to guide transfusion decisions in patients with CHF. Guidelines also recommend that pRBC transfusion decisions consider the overall clinical context rather than hemoglobin concentration alone.^24^ However, how context is meant to be incorporated into pRBC transfusion decision-making is unclear. Our results help to inform these knowledge gaps in several ways. Readily observed clinical characteristics identified in our study were associated with increased transfusion risks and may have implications for clinical decision-making. For example, a clinician caring for a patient several days into admission with hyperkalemia and low glomerular filtration rate may more strongly consider withholding pRBC transfusion for Hemoglobin concentrations of 6.5-6.9 g/dL unless clear signs of adverse effects from anemia are present. In contrast, clinicians may more strongly consider transfusion earlier in a hospitalization and in patients with low bicarbonate concentrations as being associated with lower risk. Our CATE model estimate may also have value embedded into electronic medical records as a real-time pRBC transfusion risk-benefit prediction score. Our findings that the average effect of pRBC transfusion is harmful in hospitalized patients with CHF aligns with results from other studies not specifically among patients with CHF^2,3,25^ and may suggest the need to reconsider overall recommendations for hemoglobin threshold-based transfusion practice. To realize the goal of context-specific transfusion personalization, future studies should seek to understand clinician preferences for interacting with CATE-based prediction scores, such as at what CATE cut-off clinicians would perceive benefits to outweigh risks and alternatives (e.g., iron supplementation, lower hemoglobin thresholds) to transfusion when risks are high.

Although associations between specific clinical characteristics and pRBC transfusion effectiveness were complex, several observations warrant additional consideration. Low white blood cell counts predicted pRBC transfusion benefit which may represent patients with bone marrow failure^26^ who might be expected to benefit from exogenous pRBC transfusion more than other anemia treatments such as iron infusion. Both hypo- and hyperkalemia were associated with pRBC transfusion harm; lysis of fragile stored blood leading to hyperkalemia is a well-documented complication from pRBC transfusion^27^ and massive transfusion can paradoxically cause hypokalemia.^28^ We hypothesize that pRBC transfusion in the these settings may worsen potassium derangements that further increase risks of arrythmia. The potential harms of pRBC transfusion identified in patients with low glomerular filtration rate may reflect patients with renal failure who are at higher risks of transfusion associated circulatory overload.^29^ Our findings that pRBC transfusion heterogeneity depended on timing relative to admission are consistent with our previous work among all-cause hospitalization using a different cohort and analytic method.^2^ We hypothesize that the observed benefits of pRBC transfusion early in hospitalization are due to improvements in oxygen delivery to hypoxic tissues during the acute resuscitative phase of illness.

Our study has limitations. We cannot exclude the presence of unmeasured confounders between the instrument and the outcome. Despite inclusion of over 30,000 patients in the derivation and validation cohorts and strong evidence of heterogenous treatment effects, 2SLS IV estimates of pRBC transfusion effectiveness within each CATE decile were imprecise, with several mean differences in HFDs crossing 0. We included patients with both acute and chronic CHF in analyses. However, acute CHF was not a strong driver of pRBC transfusion heterogeneity and thus it is unlikely that this decision impacted findings. The observed effects are valid in patients with hemoglobin concentrations close to 7.0 g/dL and should not be generalized to other settings.

## Conclusion

In a multicenter observational study of adult inpatients with CHF and anemia, associations between pRBC transfusion and outcomes were heterogeneous based on readily observed clinical characteristics. These results provide evidence that supports the implementation of personalized pRBC transfusion decision-making in patients with CHF and anemia.

## Data Availability

The analytic dataset for Aim 1 is not able to be shared publically due to data sharing agreement restrictions between Boston University and Premier Inc. However, we will include in the R Markdown file for Aim 1 the code required to recapitulate the analytic dataset using the raw Premier PINC AI Healthcare database. Thus, researchers seeking to replicate our results can acquire the Premier PINC AI Healthcare database from Premier Inc. separately and can use the RMarkdown file to recreate the analytic dataset in its entirety.

https://osf.io/8prh7/overview?view_only=ea9df5bbda844fa9aa86f046a5b68341

## Abbreviations

CHF: congestive heart failure
pRBC: packed red blood cell
PHD: Premier Healthcare Database
THE: heterogeneity of treatment effect
HFD: Hospital free days
SMD: absolute standardized mean differences
CATE: conditional average treatment effect
SHAP: Shapley Additive exPlanations

## Acknowledgements

This manuscript and online-only supplement were edited for clarity from January 2^nd^ 2026 to February 16^th^ 2026 with the assistance of Anthropic Claude 4.5 Haiku (claude-haiku-4-5-20251001) within the Boston University TerrierGPT platform.^30^

## Notes

Source of support: This study was funded by the American Heart Association (AHA) (24CDA1267699), and The Boston University Chobanian & Avedisian School of Medicine Department of Medicine Career Investment Award.

### Competing Interest Statement

The authors have declared no competing interest.

### Funding Statement

This study was funded by the American Heart Association (AHA) (24CDA1267699), and The Boston University Chobanian & Avedisian School of Medicine Department of Medicine Career Investment Award.

### Author Declarations

This study was deemed not human subjects research (Boston University Institutional Review Board H-44912).

